# Deep-Learning-based Quantification of Epicardial Adipose Tissue by 3D Dixon Cardiovascular Magnetic Resonance

**DOI:** 10.64898/2026.06.30.26356920

**Authors:** Halil Noyan, Richard Hickstein, Clemens Ammann, Johanna Kuhnt, Maximilian Fenski, Claudia Prieto, René M. Botnar, Thomas Hadler, Christian Hickstein, Elias Daud, Edyta Blaszczyk, Jan Gröschel, Christian Lim, Jeanette Schulz-Menger

**Affiliations:** Working Group on Cardiovascular Magnetic Resonance, Experimental and Clinical Research Center, a joint cooperation between the Charité – Universitätsmedizin Berlin and the Max-Delbrück-Center for Molecular Medicine, Berlin, Germany; DZHK (German Centre for Cardiovascular Research), partner site Berlin, Berlin, Germany; Technische Universität Berlin, Faculty IV – Electrical Engineering and Computer Science, Institute of Software Engineering and Theoretical Computer Science, Straße des 17. Juni 135, 10623 Berlin, Germany; Department of Cardiology and Nephrology, HELIOS Hospital Berlin-Buch, Berlin, Germany; Deutsches Herzzentrum der Charité, Department of Cardiology, Angiology and Intensive Care Medicine, Chariteplatz 1, Berlin, Germany; School of Biomedical Engineering and Imaging Sciences, King’s College London, London, UK; Pontificia Universidad Católica de Chile, School of Engineering and Institute for Biological and Medical Engineering, Santiago, Chile; Millennium Institute iHEALTH, Santiago, Chile

**Keywords:** Cardiovascular Magnetic Resonance, Deep-Learning, Epicardial Adipose Tissue, 3D Dixon, nnU-Net, Automated Segmentation

## Abstract

**Background:** Epicardial adipose tissue (EAT) is a metabolically active fat depot adjacent to the myocardium and the coronary arteries that can be non-invasively assessed by cardiovascular magnetic resonance (CMR). Increased EAT volume quantified by CMR has been linked to adverse cardiac remodeling, atrial fibrillation, coronary artery disease, and heart failure. Among CMR techniques, isotropic three-dimensional (3D) Dixon imaging at 1.3 x 1.3 x 1.3 mm^3^ resolution was developed to improve tissue characterization, providing fat-water signal separation for precise volumetric EAT assessment. However, manual segmentation of 3D datasets is highly time-consuming. For integration into clinical and research CMR workflows, reliable and fast automated segmentation is needed.

**Purpose:** To develop and evaluate an automated deep-learning-based pipeline for ventricular EAT quantification based on isotropic 3D Dixon CMR acquisitions.

**Methods:** An nnU-Net model was trained on 165 3D Dixon CMR cases encompassing healthy individuals and patients with underlying cardiovascular disease. The model was trained using all four Dixon phase images (opposed-phase, in-phase, fat-phase, water-phase). Manual 3D ventricular EAT segmentations served as the ground truth for training and evaluation. Performance was evaluated in 30 independent cases using Dice similarity coefficient (DSC), 95th percentile Hausdorff distance (HD95), volumetric agreement, Pearson correlation, intraclass correlation (ICC), and Bland–Altman analysis. Model performance was benchmarked against interobserver and intraobserver variability.

**Results:** Automated segmentation achieved a mean DSC of 0.896 ± 0.039 and HD95 of 1.84 ± 0.93 mm versus ground truth. Volumetric agreement with ground truth was high (r = 0.984, ICC = 0.988, p < 0.001; mean bias −0.70 mL, limits of agreement (LoA) [−10.31, 8.90] mL), exceeding interobserver agreement (bias −25.24 mL, LoA [−42.81, −7.66] mL) and comparable to intraobserver reproducibility (bias 2.72 mL, LoA [−8.73, 14.17] mL). Automated segmentation required less than one minute per case compared to 58.4 ± 7.9 minutes for manual segmentation. Two of 30 cases (6.7%) required minor manual correction, both less than five minutes.

**Conclusion:** Fully automated nnU-Net–based ventricular EAT segmentation from isotropic 3D Dixon CMR achieves accuracy comparable to intraobserver reproducibility while significantly reducing post-processing time. The approach may facilitate large-scale and longitudinal EAT quantification in CMR-based research workflows.

## Introduction

Epicardial adipose tissue (EAT) refers to the fat depot located between the myocardium and the visceral layer of the pericardium. It is in direct contact with both the myocardium and the coronary vessels without any separating tissue [1].

Previous studies quantifying EAT volume by cardiovascular magnetic resonance (CMR) have demonstrated that EAT independently predicts major adverse cardiovascular events and adds incremental prognostic value beyond established clinical and imaging parameters [2,3]. Higher EAT volume has also been linked to adverse cardiac remodeling and functional impairment [4]. These findings support the potential of EAT as an essential imaging biomarker for improved cardiovascular phenotyping and risk stratification.

CMR allows standardized and reproducible evaluation of cardiac morphology, function, and myocardial tissue characteristics, supporting comprehensive cardiac phenotyping within a single examination [5]. Despite these technical advantages, there is no standard for EAT assessment using CMR. Variability exists with respect to sequence selection, spatial resolution, anatomical definitions, and post-processing strategies [6]. These challenges are further amplified by the thin and spatially heterogeneous distribution of EAT, which makes it susceptible to partial volume effects, especially in two-dimensional acquisitions with limited through-plane resolution [7].

Among CMR techniques, 1.3 x 1.3 x 1.3 mm^3^ isotropic three-dimensional (3D) Dixon imaging enables robust differentiation between fat and water tissue signals within and around the heart. When contrast agent is applied, Late Gadolinium Enhancement (LGE) can simultaneously be assessed during the same acquisition and within the same slice location, further enhancing tissue characterization [8]. 3D Dixon CMR provides multiple phases, including opposed-phase (OP), in-phase (IP), fat phase (FP), and water phase (WP) images. In particular, the technique yields a fat phase with isotropic 3D resolution. This has the potential for precise volumetric assessment, as it permits individualized 3D reformatting and navigation, allowing anatomical boundary structures to be cross-referenced and confirmed across standardized anatomical planes, multiple planes and phases [9].

However, broad adoption of 3D Dixon CMR for EAT quantification is limited by its post-processing burden. To preserve the technique’s anatomical precision, ventricular EAT is segmented manually slice by slice across the isotropic volume, typically encompassing 80–100 slices, which can take up to one hour per case [10]. Consequently, manual 3D segmentation of ventricular EAT is not feasible for routine clinical application and large-scale studies.

Therefore, the aim of this study was to develop a fully automated deep learning–based pipeline for ventricular EAT segmentation using 1.3 x 1.3 x 1.3 mm^3^ isotropic 3D Dixon CMR and to evaluate this pipeline across a wide range of EAT volumes, including healthy individuals and patients with underlying cardiovascular disease. We hypothesized that automated 3D segmentation would achieve agreement comparable to human interobserver variability, while substantially reducing post-processing time.

## Methods

### Study Population and Dataset Splits

CMR examinations performed at our site between April 2021 and December 2025 that included non-contrast enhanced 3D Dixon and 3D Dixon Late Gadolinium Enhancement (LGE) imaging were screened retrospectively. Images of non-diagnostic quality were excluded. All source studies received approval from the local institutional ethics committee and were conducted in accordance with the Declaration of Helsinki. Written informed consent was obtained from all participants prior to inclusion.

Separation between training and test datasets was performed at the subject level, such that all images from a given individual were assigned to either the training or the test dataset, preventing data leakage between splits.

The training dataset (n = 165) was assembled to cover the key sources of variability relevant for automated EAT segmentation. It comprised both healthy individuals with normal cardiac structure and function and patients with underlying cardiovascular disease, spanning a wide range of EAT volumes from low to high burden (Figure 1).

**Figure 1.**
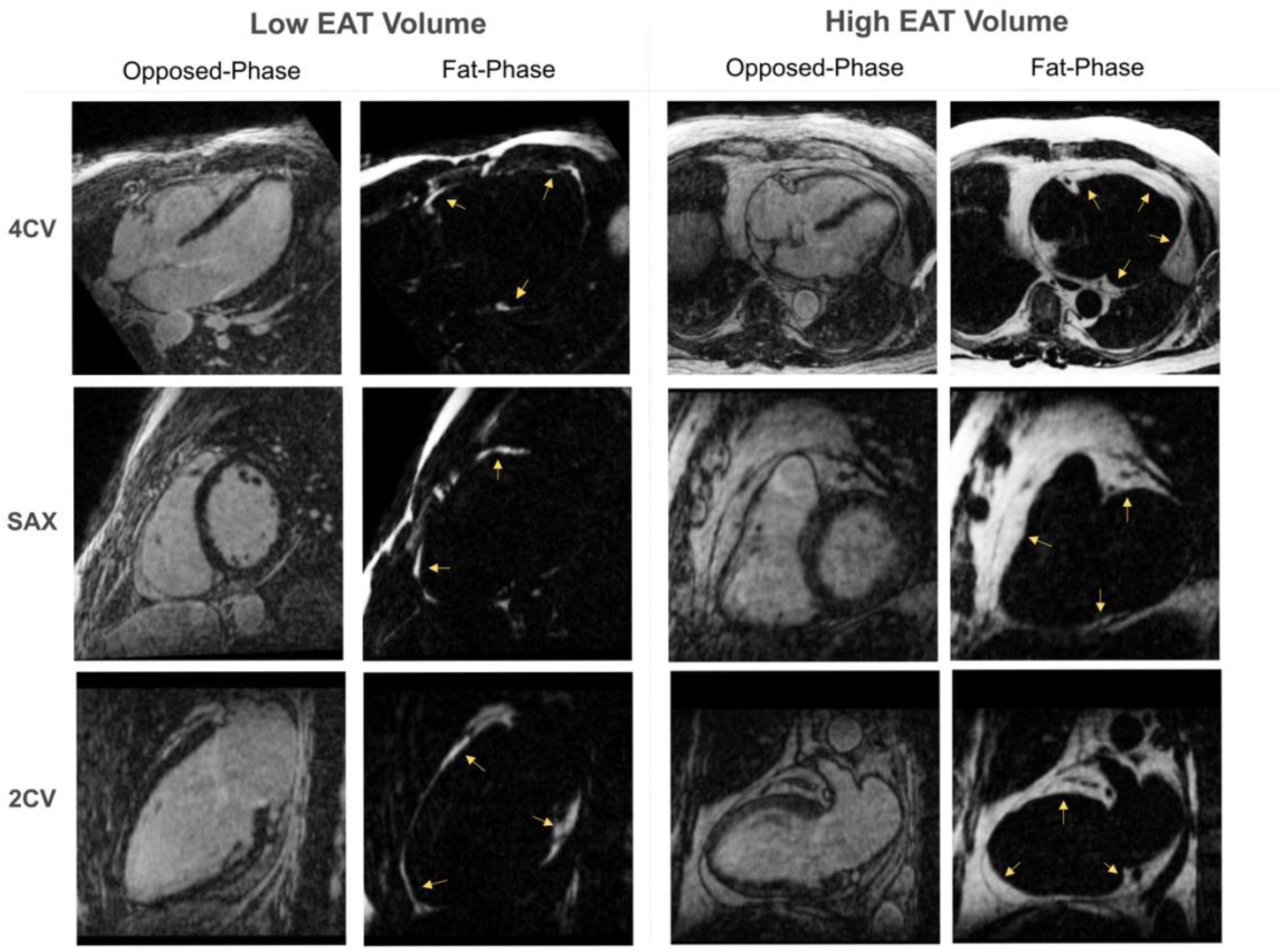
Low (left) and high (right) ventricular epicardial adipose tissue (EAT) burden on 1.3 x 1.3 x 1.3 mm³ isotropic 3D Dixon CMR, shown in opposed-phase and fat-phase images across four-chamber (4CV), short-axis (SAX), and two-chamber (2CV) views. EAT appears as hyperintense signal on fat-phase images (yellow arrows).

The cohort was further enriched for pathological features to allow systematic evaluation of their potential impact on automated EAT segmentation: Intramyocardial fat may blur the epicardial boundary, as it shares signal characteristics with EAT on Dixon fat-phase images. Myocardial scar manifesting as LGE may pose an additional challenge, as gadolinium-induced alterations in local signal intensity in affected regions can disrupt the anatomical contrast between myocardium and adjacent EAT across the 3D Dixon CMR phases used for automated segmentation. In addition, the cohort covered a range of image quality grades, reflecting the variability in signal-to-noise ratio and artifact burden encountered in routine clinical acquisitions. Physician-assigned clinical diagnoses contributing to the cohort’s composition spanned cardiomyopathies including myocardial involvement in systemic disorders as well as coronary artery disease. A detailed overview of the diagnoses is provided in Supplementary Table 1.

A separate test dataset (n = 30) was defined a priori to provide an independent and completely unseen dataset for performance evaluation across the variability dimensions described above, while preserving a sufficiently large proportion of the data for model training. The test dataset was sampled using a simulated annealing approach with temperature set to zero, thus acting as a greedy randomized local search [11]. A multi-objective cost function was used to ensure a wide range of EAT volumes in both dataset splits, balanced representation of cardiac conditions, a normal distribution of image quality, and equal gender proportion within the test dataset.

### CMR Acquisition

CMR examinations were performed on clinical 1.5 Tesla (T) (MAGNETOM Avanto Fit, Siemens Healthineers AG, Forchheim, Germany, n = 182) and 3 T (MAGNETOM Skyra Fit, Siemens Healthineers AG, Forchheim, Germany, n = 13) scanners. Of the 195 patients, 178 received a gadolinium-based contrast agent, while 17 underwent non-contrast enhanced imaging only. Contrast media dose was 0.2 mmol/kg Gadoteridol (ProHance, Bracco, Milan, Italy) or 0.15 mmol/kg Gadobutrol (Gadovist, Bayer Healthcare, Berlin, Germany) in 1.5T and in 3T, respectively.

Non-contrast enhanced 3D Dixon and the 3D Dixon LGE images were acquired using a prototype spoiled gradient-echo sequence with inversion recovery preparation, two-point Dixon encoding, and compressed sensing. Imaging was performed in the transverse plane during free breathing with ECG gating and whole-heart coverage. The acquisition and reconstruction of this sequence have been described and characterized in detail previously [8,12]. Each acquisition produced four 3D image datasets, comprising OP, IP, FP, and WP images [13].

The non-contrast enhanced 3D Dixon sequence was performed at 1.5 T with an isotropic resolution of 1.2 × 1.2 × 1.2 mm³, echo times TE1 of 2.38 ms and TE2 of 4.67 ms, flip angle of 15°, receiver bandwidth of 497 Hz per pixel, and a field of view of 320 × 320 mm².

The 3D Dixon LGE sequence was acquired at both field strengths with an isotropic resolution of 1.3 × 1.3 × 1.3 mm³. At 1.5 T, LGE parameters comprised a flip angle of 20°, receiver bandwidth of 495 Hz per pixel, and a field of view of 311 × 311 mm². At 3 T, all patients received intravenous contrast agent, and a post-contrast Dixon sequence was acquired with echo times TE1 of 1.31 ms and TE2 of 2.81 ms, a flip angle of 15°, a receiver bandwidth of 650 Hz per pixel, and a field of view of 399 × 399 mm².

### Image Quality Assessment

Image quality was assessed using a four-point Likert scale, ranging from 0 to 3, as previously published by our working group [12]. Grade 0 indicated non-diagnostic image quality; grade 1, fair quality with potentially impaired diagnostic confidence; grade 2, adequate image quality with minor artifacts; and grade 3, excellent image quality without relevant artifacts. Image quality was assessed by HN prior to segmentation. Cases rated as grade 0 were excluded from the training and test datasets.

### Manual Ground Truth Segmentation

Manual segmentation of ventricular EAT was performed using the open-source software 3D Slicer (version 5.8.1) [14]. All manual segmentations served as ground truth and were performed by observer HN. The segmentations for interobserver analysis were performed by a second independent observer (CL) blinded to the results of the initial ground truth segmentation. Segmentation was performed primarily in the short-axis orientation. Long-axis views (two-chamber and four-chamber) were used as cross-references to confirm anatomical boundaries, particularly in apical regions where EAT delineation is challenging when relying solely on short-axis images [5].

The most basal slice included in the ventricular EAT segmentation was defined in accordance with post-processing guidelines for short axis ventricular functional analysis, ensuring that epicardial fat surrounding the left ventricular outflow tract and atria was excluded [5]. An example of the most basal slice is shown in Figure 2. EAT volume was quantified by multiplying voxel size with the total number of segmented voxels.

**Figure 2.**
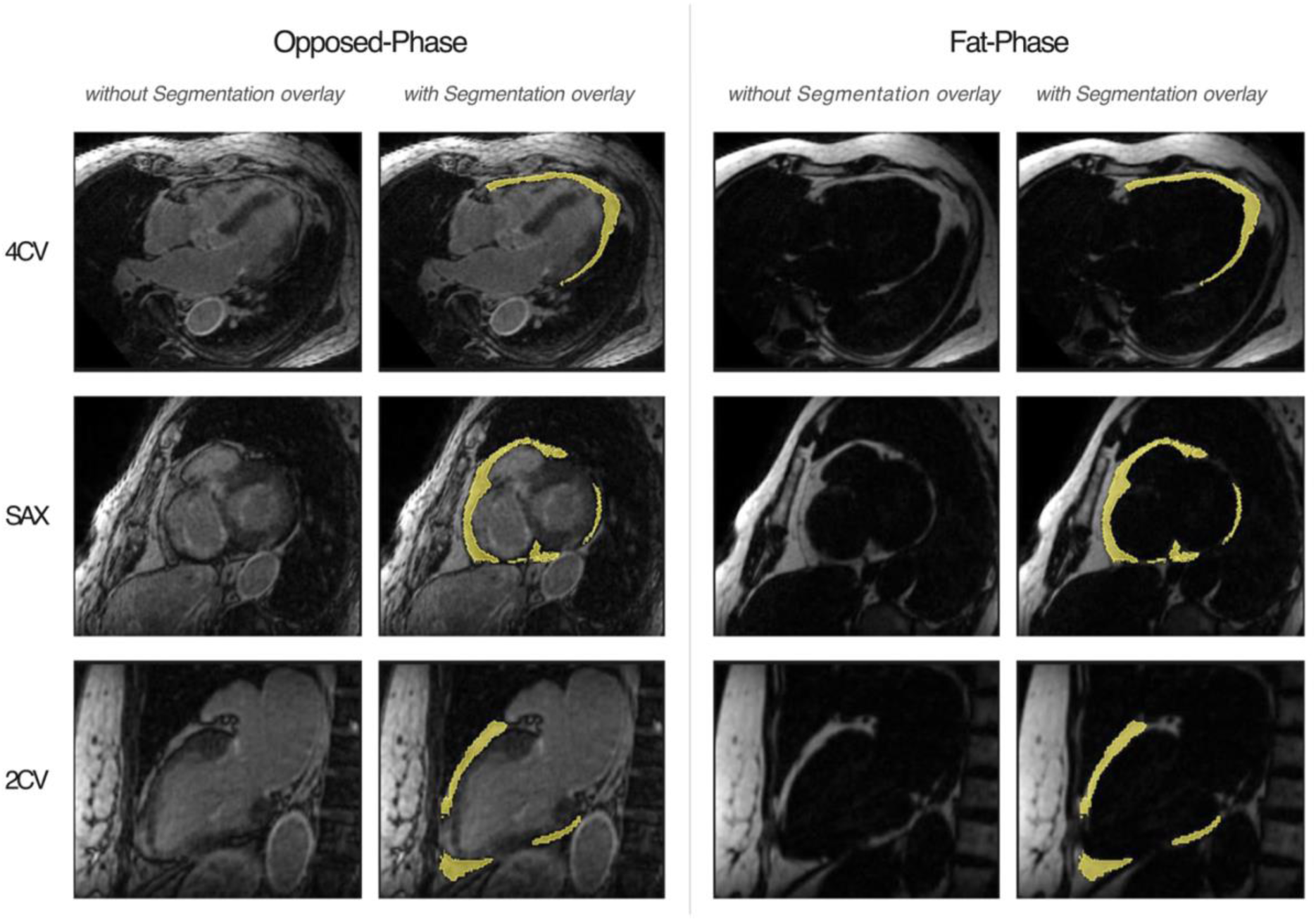
Representative 3D Dixon LGE CMR images in four-chamber (4CV), short-axis (SAX), and two-chamber (2CV) views, shown in opposed-phase (left) and fat-phase (right), without and with ventricular EAT segmentation overlay. Epicardial adipose tissue is highlighted in yellow. SAX slices correspond to the most basal ventricular level included in the segmentation.

### Model Architecture and Training

A 3D nnU-Net model (nnUNetv2, Version 2.6.2) [15] was trained to enable automated EAT segmentation (Figure 3). The nnU-Net framework was selected due to its established benchmark performance in biomedical image segmentation and its ability to automatically configure preprocessing steps, network topology, and training hyperparameters based on dataset-specific characteristics. Detailed architecture and training parameters are provided in the Supplementary Material.

**Figure 3.**
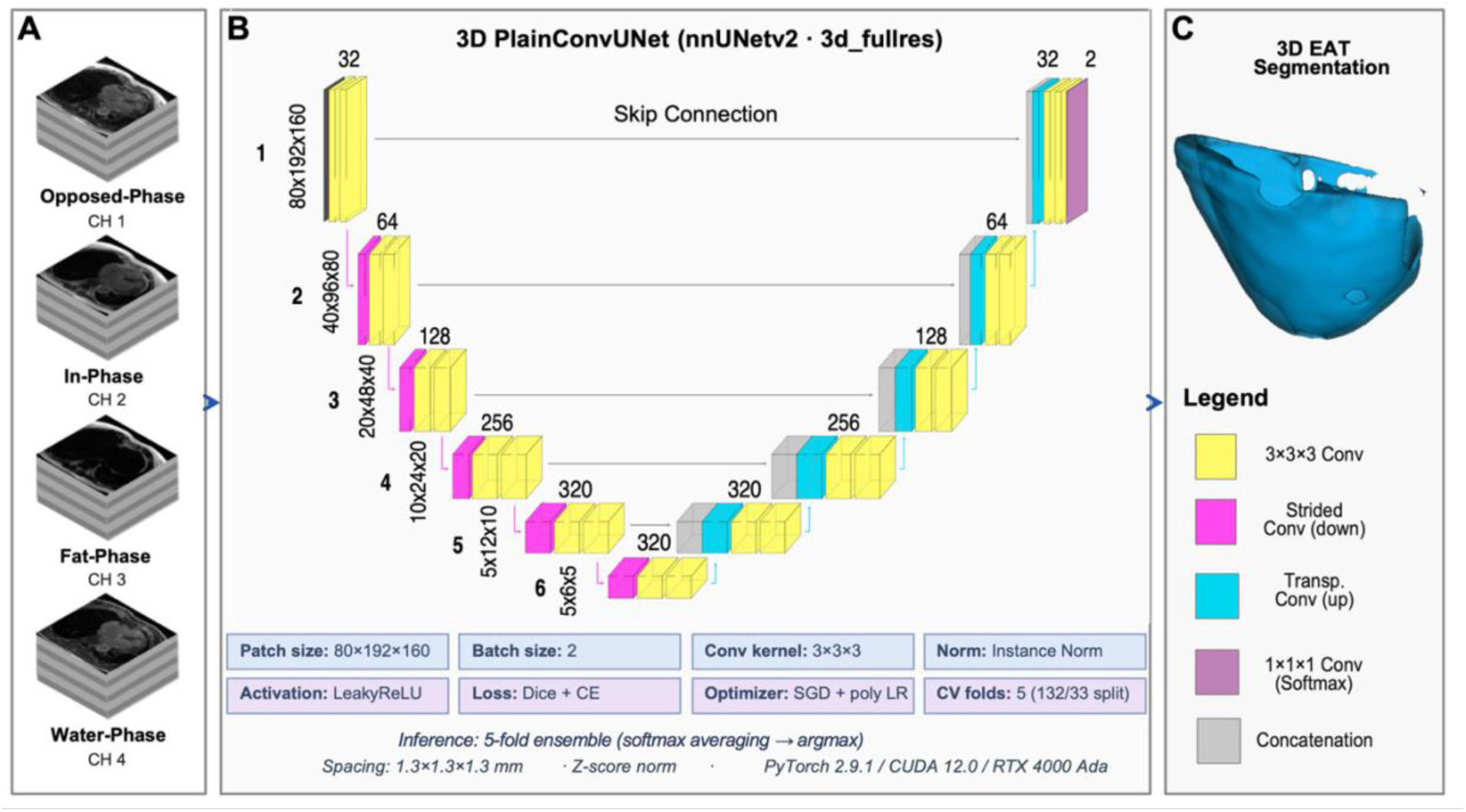
(A) Four-channel 3D Dixon CMR input (opposed-, in-, fat-, water-phase). (B) 3D nnU-Net architecture for automated EAT segmentation. (C) Manually segmented 3D ventricular EAT volume (ground truth). CMR, cardiac magnetic resonance; EAT, epicardial adipose tissue; CH, channel.

The network received four input channels corresponding to the OP, IP, FP and WP images of the 3D Dixon CMR sequence. Manual 3D ventricular EAT segmentations served as ground truth.

### Performance Evaluation

Qualitative evaluation focused on identifying major segmentation errors, including anatomically implausible segmentations, gross over-segmentation, or missed EAT regions, particularly in basal and apical slices.

Quantitative assessment was performed by comparing model performance against human interobserver variability and intraobserver variability. Interobserver variability was assessed using a separate unseen dataset of 30 cases not used for training. The intraobserver variability was assessed by having one observer independently segment the same cases twice, separated by a blanking period of two months. Spatial overlap was assessed using the Dice similarity coefficient (DSC), boundary accuracy using the 95th percentile Hausdorff distance (HD95), and volumetric agreement using absolute and relative volume differences. Pearson correlation coefficients, intraclass correlation coefficients (ICC), and Bland–Altman analysis were used to assess agreement in EAT volume estimation.

In the subgroup analysis the model robustness was further evaluated by assessing segmentation performance separately across the variability dimensions of the study cohort, using DSC and absolute volume difference against ground truth. Performance was compared between healthy individuals and patients, and between cases with low and high EAT burden, separated at the cohort median. In addition, performance was assessed in subgroups defined by the presence or absence of LGE, the presence or absence of myocardial fat infiltration and across image quality grades 1 to 3.

### Segmentation Efficiency and Correction Workload

To estimate the proportion of automated segmentations potentially requiring manual correction, a dual-criterion approach was applied [16]. Tolerance thresholds were derived from the intraobserver variability analysis. A volume threshold was defined as 1.96 standard deviations of the absolute intraobserver volume difference (threshold: 7.02 mL). A DSC threshold was defined as the median intraobserver DSC minus the median absolute deviation (threshold: 0.865). An automated segmentation was classified as potentially requiring manual correction if both criteria were simultaneously met. This combined criterion captures clinically relevant segmentation deviations while avoiding flagging cases with minor boundary differences but adequate volumetric agreement, or cases with larger volume differences but high spatial overlap. Manual segmentation was timed prospectively during ground truth annotation. Automated inference time was recorded for all test cases.

## Statistical Analysis

Statistical analysis was performed using Python (Version 3.12.9) with the following libraries: NumPy (Version 2.3.1) for numerical computations, pandas (Version 2.3.1) for data handling and tabular processing, SciPy (Version 1.16.1) for statistical testing, and pynrrd (Version 1.1.3) for processing nearly raw raster segmentation data (NRRD) [17–20]. The significance threshold was set to p=0.05. Continuous variables are reported as mean ± standard deviation (SD) or median [interquartile range (IQR)], as appropriate.

## Results

### Study Population and Dataset Characteristics

A total of 206 CMR examinations were identified, of which 195 were subsequently included in the final analysis (the training dataset, n = 165 and test dataset, n = 30). Eleven examinations were excluded due to non-diagnostic image quality. Demographic characteristics, image quality distribution, and contrast agent administration rates were comparable between both datasets and are summarized in Table 1.

**Table 1:**
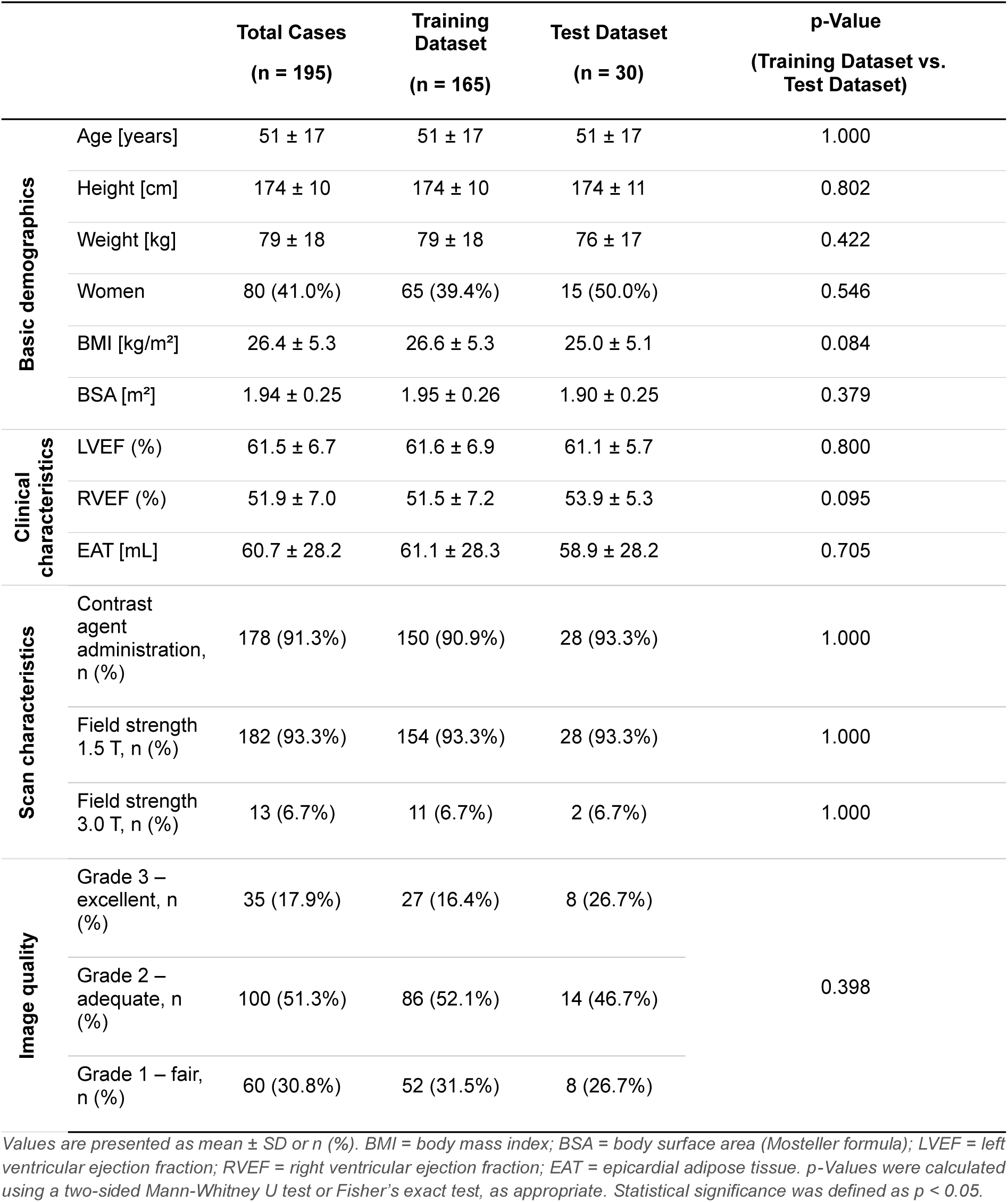
Characteristics of the study population by dataset.

|  |  | **Total Cases**<br>**(n = 195)** | **Training**<br>**Dataset**<br>**(n = 165)** | **Test Dataset**<br>**(n = 30)** | **p-Value**<br>**(Training Dataset vs.**<br>**Test Dataset)** |
| --- | --- | --- | --- | --- | --- |
| **Basic demographics** | Age [years] | 51 ± 17 | 51 ± 17 | 51 ± 17 | 1.000 |
|  | Height [cm] | 174 ± 10 | 174 ± 10 | 174 ± 11 | 0.802 |
|  | Weight [kg] | 79 ± 18 | 79 ± 18 | 76 ± 17 | 0.422 |
|  | Women | 80 (41.0%) | 65 (39.4%) | 15 (50.0%) | 0.546 |
|  | BMI [kg/m<sup>2</sup>] | 26.4 ± 5.3 | 26.6 ± 5.3 | 25.0 ± 5.1 | 0.084 |
|  | BSA [m<sup>2</sup>] | 1.94 ± 0.25 | 1.95 ± 0.26 | 1.90 ± 0.25 | 0.379 |
| **Clinical**<br>**characteristics** | LVEF (%) | 61.5 ± 6.7 | 61.6 ± 6.9 | 61.1 ± 5.7 | 0.800 |
|  | RVEF (%) | 51.9 ± 7.0 | 51.5 ± 7.2 | 53.9 ± 5.3 | 0.095 |
|  | EAT [mL] | 60.7 ± 28.2 | 61.1 ± 28.3 | 58.9 ± 28.2 | 0.705 |
| **Scan characteristics** | Contrast agent administration, n (%) | 178 (91.3%) | 150 (90.9%) | 28 (93.3%) | 1.000 |
|  | Field strength 1.5 T, n (%) | 182 (93.3%) | 154 (93.3%) | 28 (93.3%) | 1.000 |
|  | Field strength 3.0 T, n (%) | 13 (6.7%) | 11 (6.7%) | 2 (6.7%) | 1.000 |
| **Image quality** | Grade 3 – excellent, n (%) | 35 (17.9%) | 27 (16.4%) | 8 (26.7%) | 0.398 |
|  | Grade 2 – adequate, n (%) | 100 (51.3%) | 86 (52.1%) | 14 (46.7%) |  |
|  | Grade 1 – fair, n (%) | 60 (30.8%) | 52 (31.5%) | 8 (26.7%) |  |

Values are presented as mean ± SD or n (%). BMI = body mass index; BSA = body surface area (Mosteller formula); LVEF = left ventricular ejection fraction; RVEF = right ventricular ejection fraction; EAT = epicardial adipose tissue. p-Values were calculated using a two-sided Mann-Whitney U test or Fisher's exact test, as appropriate. Statistical significance was defined as $p < 0.05$ .

EAT volume distribution was comparable between the training and test set, covering the full spectrum of EAT volumes observed in the overall dataset (p=0.705, Figure 4).

**Figure 4.**
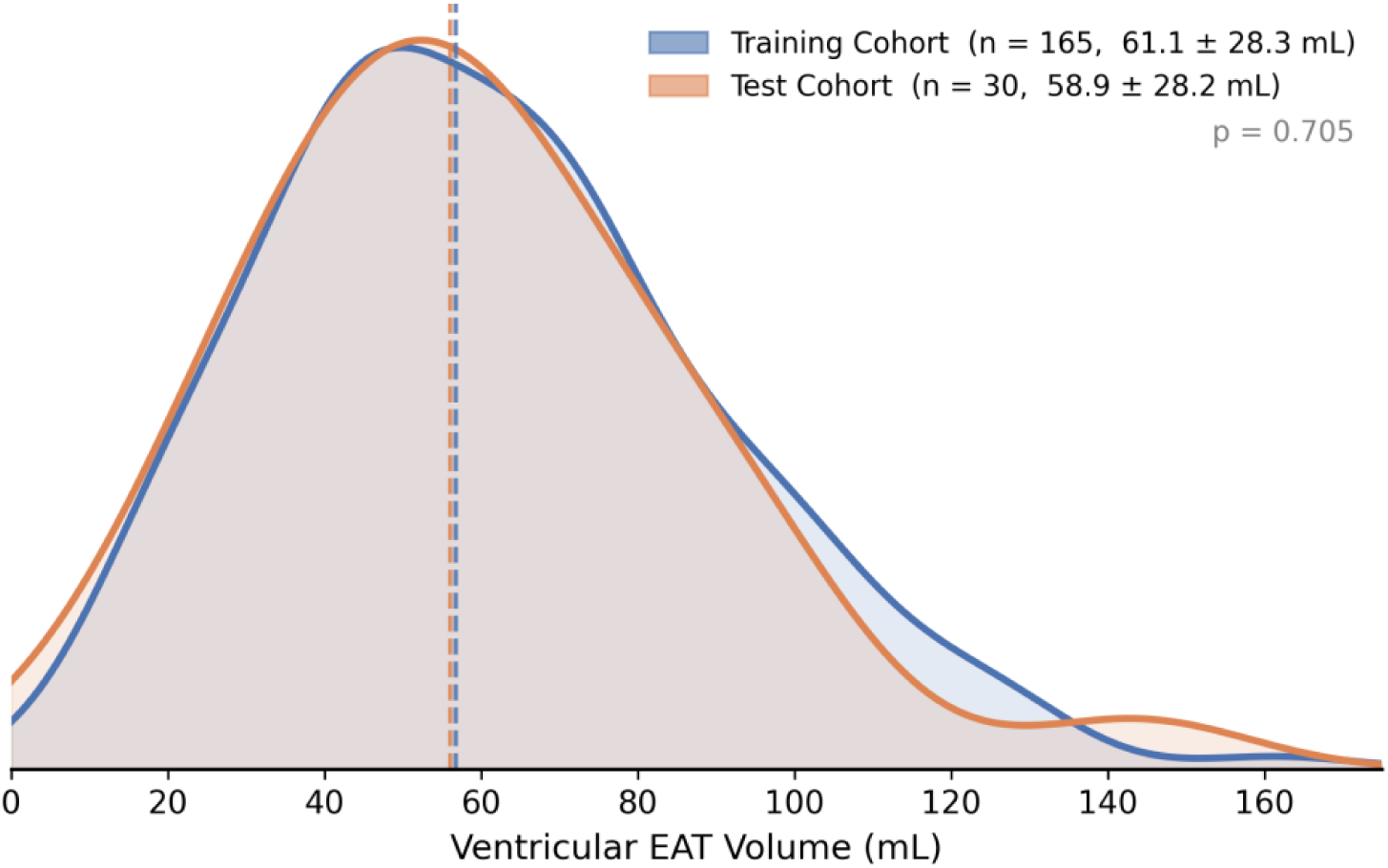
Ventricular epicardial adipose tissue (EAT) volume distribution in the training and test dataset. Kernel density estimate (KDE) curves with shaded areas are shown for the training dataset (blue) and the test dataset (orange). Dashed vertical lines indicate the respective dataset medians.

### Model Evaluation

Mean manual segmentation time was 58.4 ± 7.9 minutes per case. Automated inference was completed in under one minute per case, representing a significant reduction in processing time. Based on the dual-criterion approach derived from intraobserver variability, 2 of 30 cases (6.7%) were flagged for potential manual correction, requiring 4:12 and 4:41 minutes of additional editing time, respectively.

### Qualitative Segmentation Analysis

Qualitative assessment revealed no anatomically implausible segmentations. No systematic failure pattern was observed in healthy individuals or patients, regardless of EAT burden, LGE, or myocardial fat infiltration. A representative case is shown in Figure 5.

**Figure 5.**
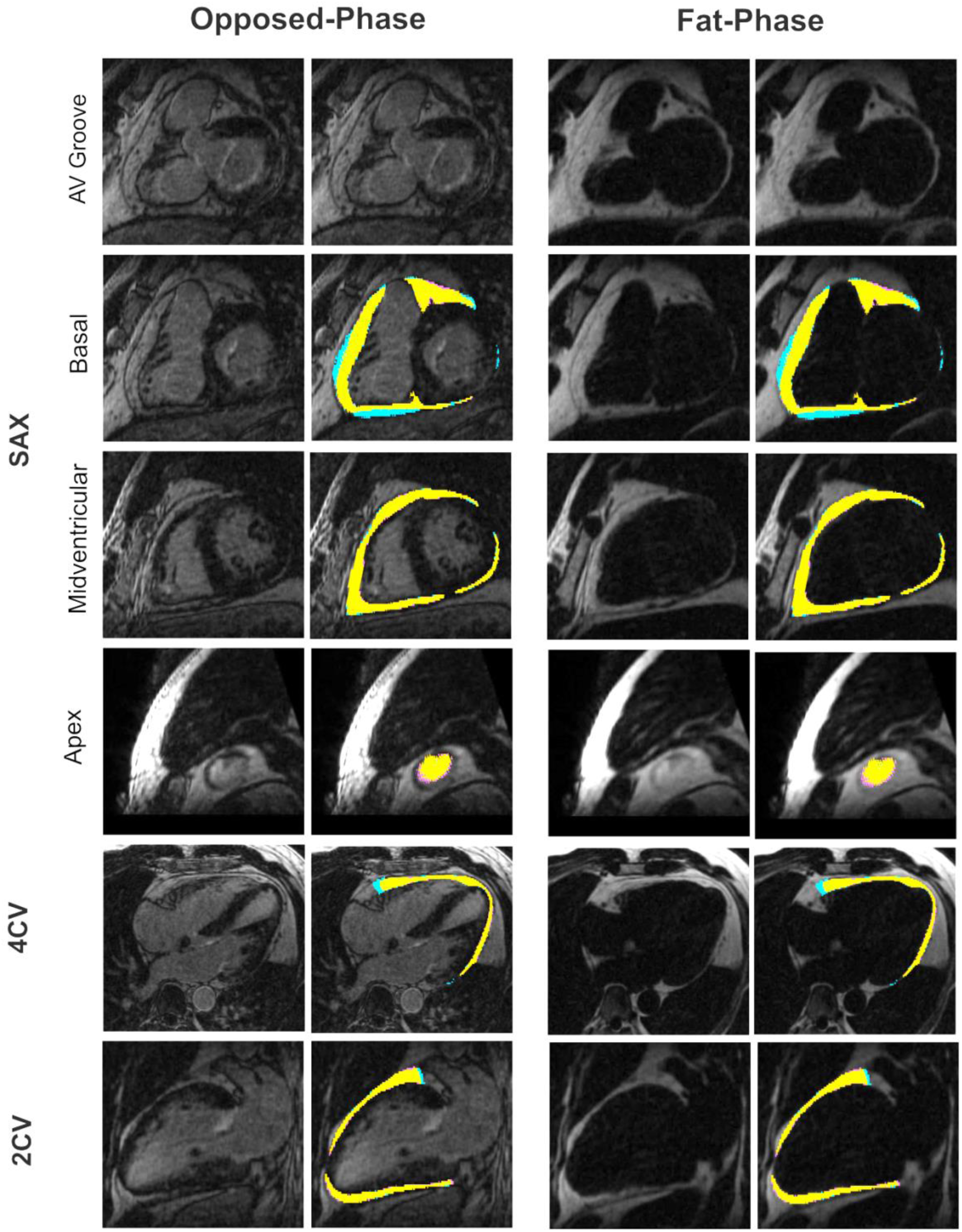
Qualitative comparison of automated EAT segmentation against manual ground truth across standard cardiac views. Each row shows a different view (SAX at atrioventricular (AV) groove, basal, midventricular, and apical levels; 4CV: four chamber view; 2CV: two chamber view) in opposed-phase and fat-phase images. Segmentation overlays (2^nd^ and 4^th^ column) are shown next to the corresponding source image (1^st^ and 3^rd^ column). Blue overlays indicate manual ground truth, magenta indicates model prediction, and orange indicates voxel-wise agreement between the two.

Three-dimensional renderings of the same representative case confirmed spatially consistent agreement (Figure 6).

**Figure 6.**
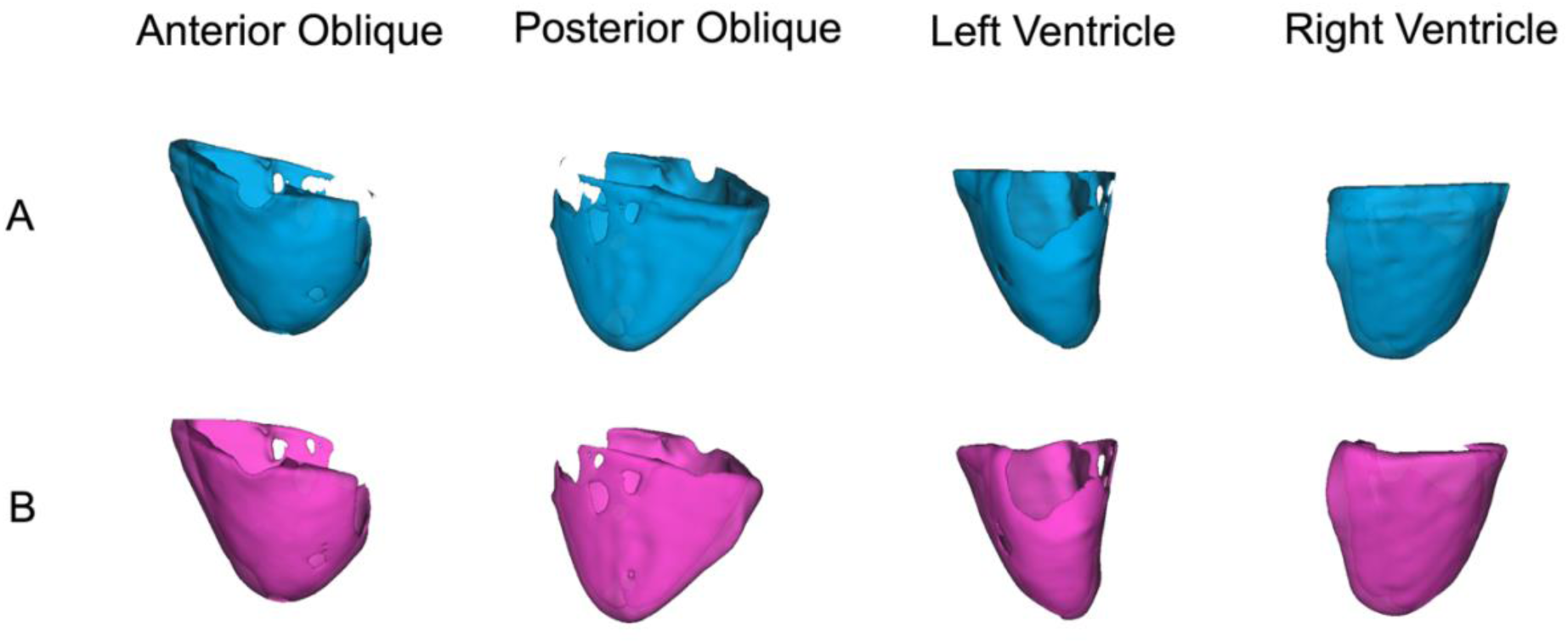
Three-dimensional renderings of epicardial adipose tissue (EAT) for a representative case, shown from four viewing angles (anterior oblique, posterior oblique, left ventricle, and right ventricle). (A) ground truth segmentation (blue); (B) model prediction (magenta).

Based on the dual-criterion approach derived from intraobserver variability, 2 of 30 cases (6.7%) were flagged for potential manual correction (Figure 7). Deviations from ground truth occurred predominantly in basal slices, where in some cases the model also segmented the atrioventricular groove EAT.

**Figure 7.**
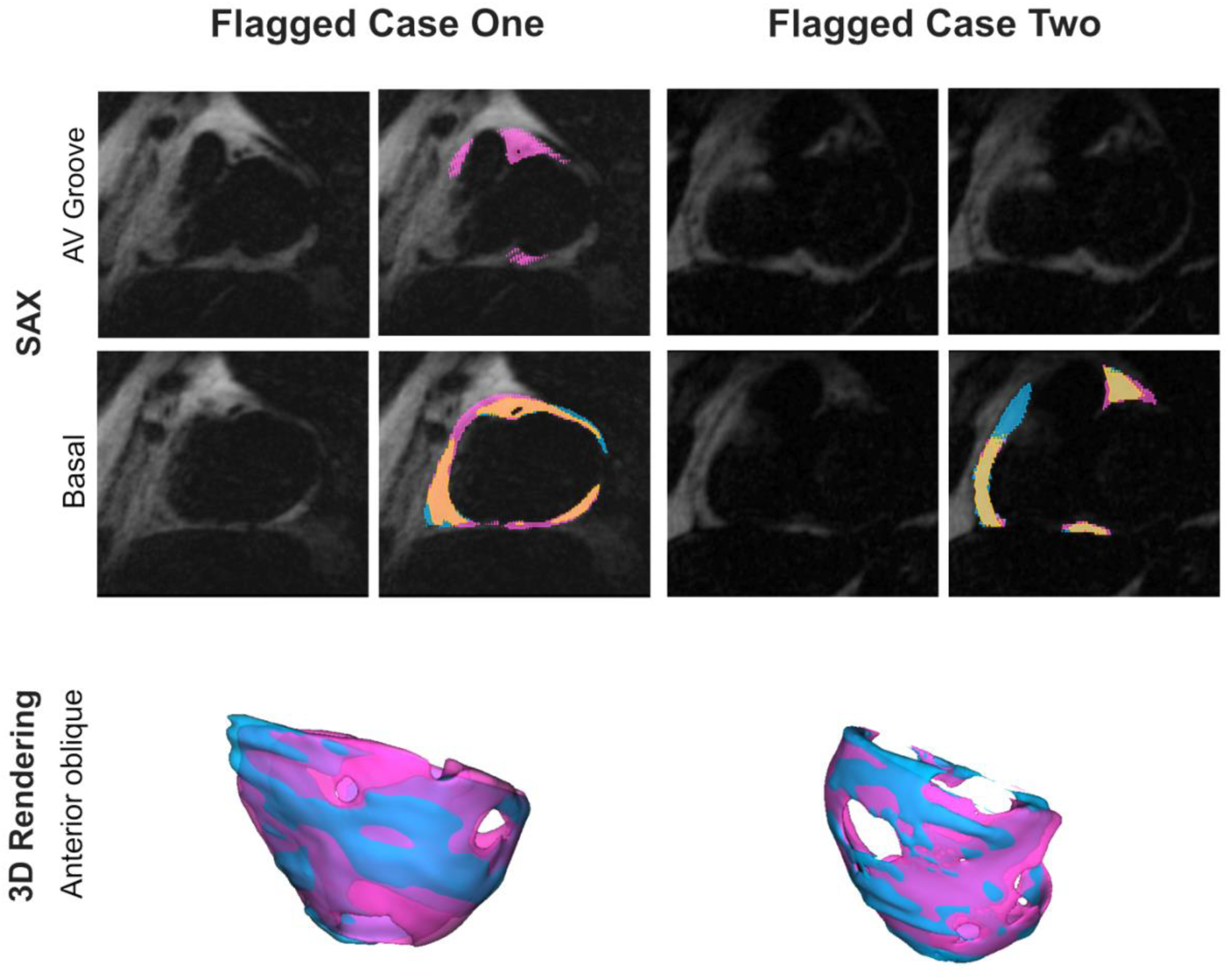
Cases flagged for potential manual correction (2/30, 6.7%). Top rows: short-axis views with source images and segmentation overlays. Bottom row: 3D renderings, anterior oblique view. Blue: ground truth; magenta: prediction; orange: agreement.

Both cases flagged for potential correction were contrast enhanced studies from patients with underlying cardiovascular disease (borderline arrhythmogenic right ventricular cardiomyopathy (ARVC) and hypertrophic cardiomyopathy (HCM), respectively). The HCM case additionally showed myocardial fat infiltration and LGE, alongside fair image quality (grade 1). Extended two-dimensional and three-dimensional visualizations of both flagged cases are provided in Supplementary Figures S1–S4.

### Interobserver Variability

Overall, interobserver agreement was good with a DSC of 0.766 ± 0.066 and HD95 of 13.05 ± 4.08 mm (95% CI: 11.60–14.54 mm), with deviations predominantly in basal slices. Volumetrically, observer CL consistently segmented larger EAT volumes than observer HN (95.54 ± 37.26 mL vs. 70.30 ± 30.00 mL), despite a strong Pearson correlation (r = 0.988, p < 0.001, ICC = 0.755, 95% CI: 0.598–0.827). Bland–Altman analysis confirmed a systematic bias of −25.24 mL (limits of agreement (LoA): −42.81 to −7.66 mL, Figure 8).

**Figure 8.**
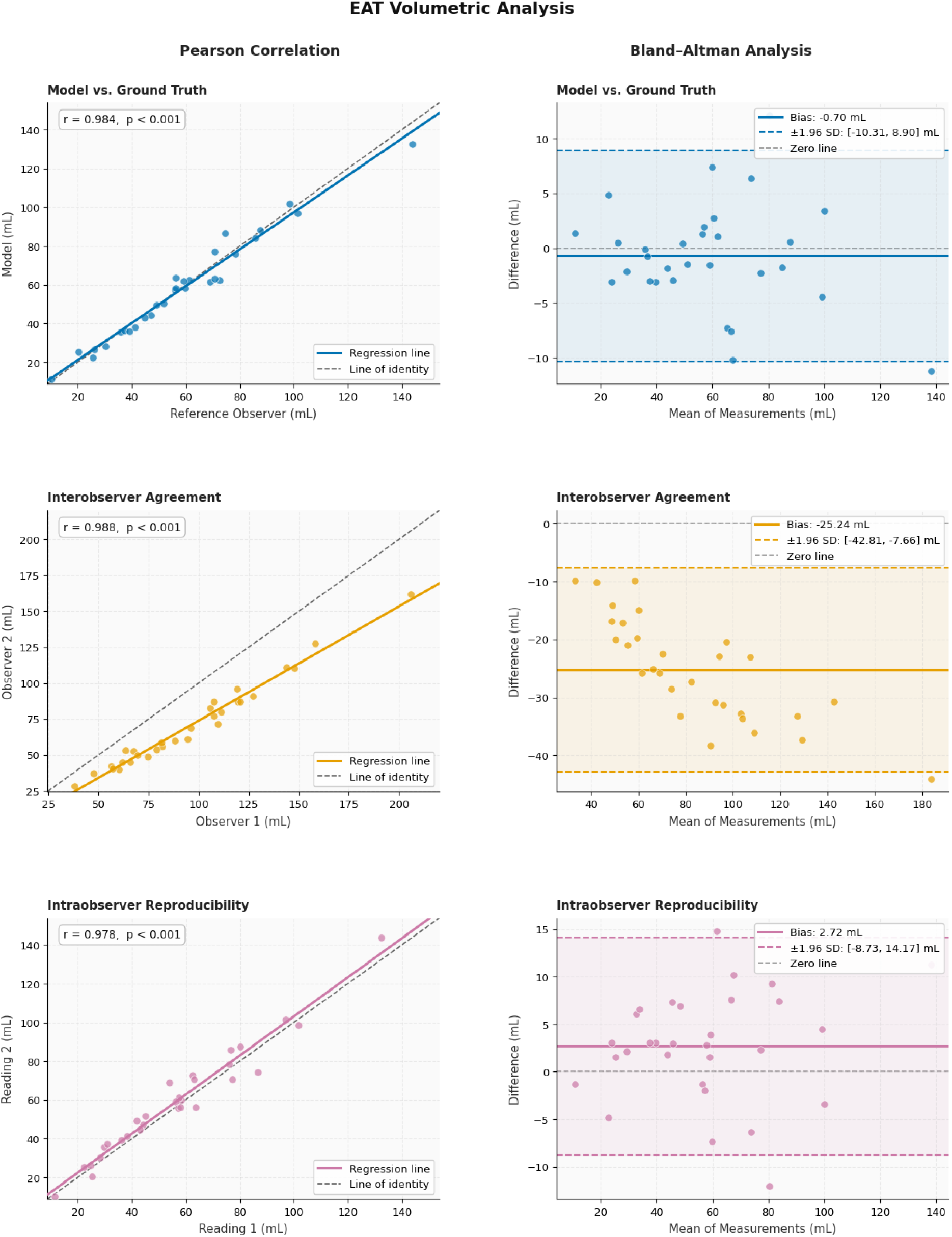
Pearson correlation (left) and Bland–Altman analysis (right) of ventricular epicardial adipose tissue (EAT) volumes across all 30 cases, shown for model versus ground truth (top), interobserver agreement (middle), and intraobserver reproducibility (bottom.)

### Intraobserver Variability

Repeated segmentations by observer HN showed high reproducibility, with a DSC of 0.885 ± 0.041 and an HD95 of 2.59 ± 1.69 mm (95% CI: 2.03–3.21 mm). Volumetric agreement was excellent (58.66 ± 27.78 mL vs. 55.94 ± 26.94 mL; absolute volume difference 5.29 ± 3.58 mL; r = 0.978, p < 0.001; ICC = 0.988). Bland–Altman analysis showed a bias of 2.72 mL (LoA: −8.73 to 14.17 mL, Figure 8).

### Model Performance

Automated segmentation outperformed interobserver agreement in spatial overlap (DSC 0.896 ± 0.039 vs. 0.766 ± 0.066, p < 0.001) and was comparable to intraobserver reproducibility (DSC 0.885 ± 0.041, p = 0.198). Likewise, HD95 was lower than for interobserver variability (1.84 ± 0.93 mm vs. 13.05 ± 4.08 mm, p < 0.001) and comparable to intraobserver variability (2.59 ± 1.69 mm, p = 0.075). Volumetrically, the absolute volume difference was substantially lower than interobserver variability (3.62 ± 3.31 mL vs. 25.24 ± 8.97 mL, p < 0.001) and slightly lower than intraobserver variability (5.29 ± 3.58 mL, p = 0.029, Table 3). Pearson correlation and Bland–Altman analysis confirmed excellent agreement between automated and ground truth segmentation (r = 0.984, p < 0.001, bias −0.70 mL, LoA: −10.31 to 8.90 mL, Figure 8).

**Table 3:**
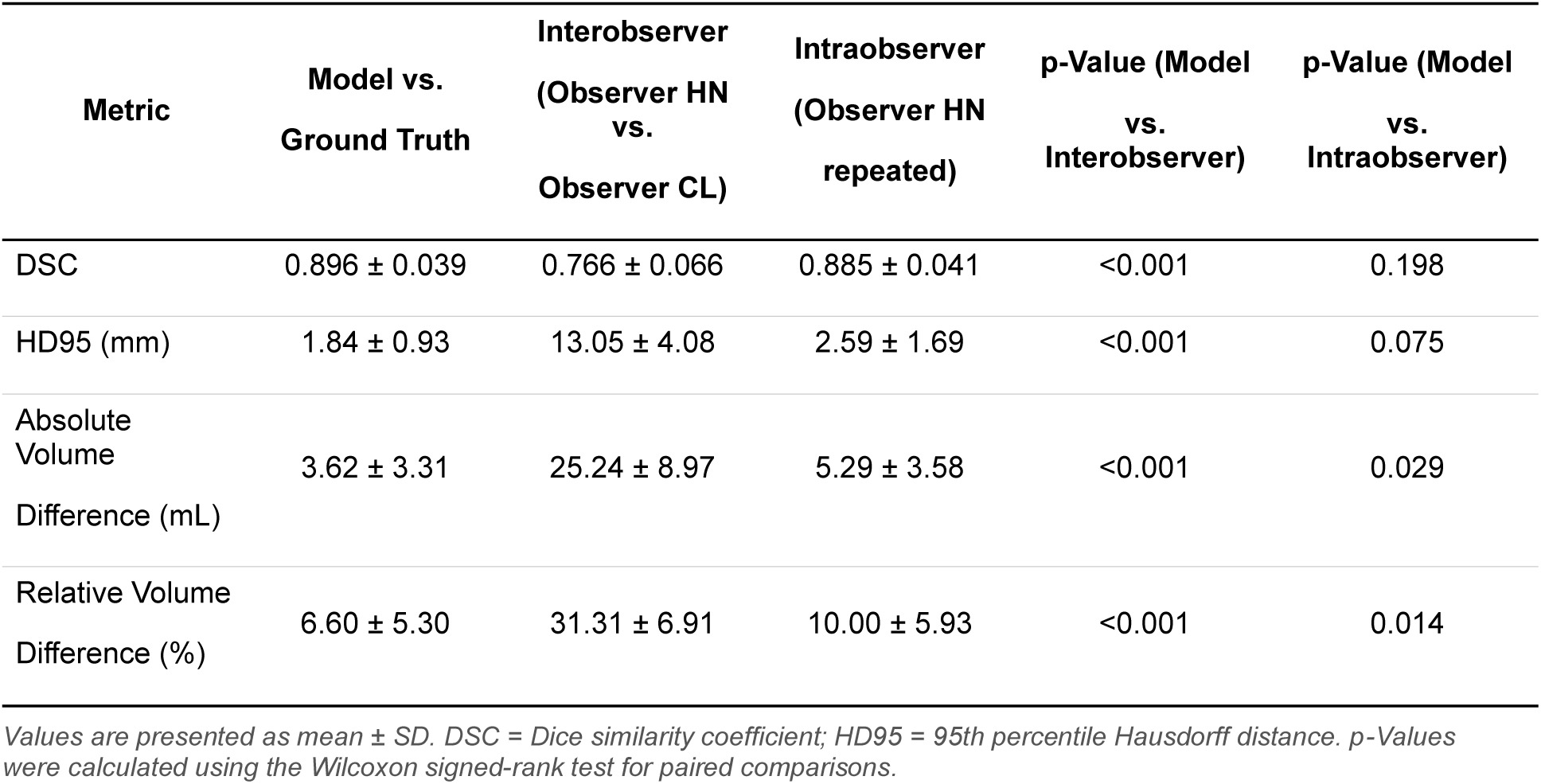
Quantitative Evaluation of Segmentation Accuracy.

| Metric | Model vs.<br>Ground Truth | Interobserver<br>(Observer HN<br>vs.<br>Observer CL) | Intraobserver<br>(Observer HN<br>repeated) | p-Value (Model<br>vs.<br>Interobserver) | p-Value (Model<br>vs.<br>Intraobserver) |
| --- | --- | --- | --- | --- | --- |
| DSC | $0.896 \pm 0.039$ | $0.766 \pm 0.066$ | $0.885 \pm 0.041$ | <0.001 | 0.198 |
| HD95 (mm) | $1.84 \pm 0.93$ | $13.05 \pm 4.08$ | $2.59 \pm 1.69$ | <0.001 | 0.075 |
| Absolute<br>Volume<br>Difference (mL) | $3.62 \pm 3.31$ | $25.24 \pm 8.97$ | $5.29 \pm 3.58$ | <0.001 | 0.029 |
| Relative Volume<br>Difference (%) | $6.60 \pm 5.30$ | $31.31 \pm 6.91$ | $10.00 \pm 5.93$ | <0.001 | 0.014 |

Values are presented as mean ± SD. DSC = Dice similarity coefficient; HD95 = 95th percentile Hausdorff distance. p-Values were calculated using the Wilcoxon signed-rank test for paired comparisons.

### Subgroup Analyses

Segmentation accuracy was slightly higher in healthy individuals (DSC 0.908 ± 0.025, absolute volume difference 1.89 ± 1.14 mL) than in patients with underlying cardiovascular disease (DSC 0.890 ± 0.046, absolute volume difference 4.79 ± 4.51 mL).

Stratification by EAT burden revealed comparable performance in both the low EAT volume group (≤56 mL EAT, DSC 0.887 ± 0.041, absolute volume difference 2.28 ± 1.92 mL) and the high EAT volume group (>56 mL EAT, DSC 0.905 ± 0.040, absolute volume difference 5.37 ± 4.86 mL).

Performance was comparable in cases with intramyocardial LGE (DSC 0.880 ± 0.060, absolute volume difference 7.54 ± 6.48 mL) and without intramyocardial LGE (DSC 0.899 ± 0.037, absolute volume difference 3.08 ± 2.91 mL). Similarly, performance was comparable in cases with myocardial fat infiltration (DSC 0.878 ± 0.059, absolute volume difference 6.51 ± 7.84 mL) and in cases without fat infiltration (DSC 0.900 ± 0.037, absolute volume difference 3.29 ± 2.62 mL). Model performance was also consistent across image quality grades (grade 3: DSC 0.912 ± 0.023; grade 2: DSC 0.898 ± 0.041; grade 1: DSC 0.876 ± 0.049).

## Discussion

In this study, a fully automated deep learning–based pipeline for ventricular EAT segmentation on isotropic 3D Dixon CMR achieved performance consistent with intraobserver reproducibility in both healthy individuals and patients, across a broad distribution of EAT burden, pathological imaging features, and image quality. Post-processing time was reduced from around an hour per case to one minute while maintaining segmentation precision.

### Model Performance

A central methodological advantage of automated EAT quantification is that it removes the systematic bias that inevitably arises between different human observers. Despite high individual precision, a second observer introduced a reproducible volumetric offset, as expected for manual segmentation of thin structures such as EAT. This matters for multi-observer and longitudinal studies, where a shift between observers or time points can be misinterpreted as a biological change. By eliminating this source of inter-observer variability, an automated model strengthens comparisons across observers, centers, and time points [21,22].

Most of the remaining deviations between automated and manual segmentation occurred in basal slices, as the anatomical transition between ventricular and atrial EAT at the atrioventricular groove provides no distinct landmark that either a human observer or a model can rely on [23]. In our case, the model’s segmentation occasionally extended past the basal boundary defined by the segmentation protocol and included atrioventricular groove fat. These deviations are not model-specific errors but reflect a challenge in basal boundary definition that is recognized in CMR standardization efforts [5].

### Subgroup Performance

As AI-based tools move toward clinical application in patients with complex cardiac disease [24], their robustness to pathological imaging features becomes essential. The performance of deep learning models often drops when presented with data differing from the distribution seen during training, a limitation referred to as distribution shift [25]. Although the model uses all four Dixon phases as input, intramyocardial fat is a particular concern, as it shares the hyperintense signal of EAT on the fat-phase images and may thus blur the epicardial boundary. Scar-related LGE and reduced image quality pose additional challenges. Despite these challenges, the model maintained consistent performance across the full range of EAT burden, in subgroups with intramyocardial LGE or fat infiltration, and across image quality grades. The comparable performance in the presence and absence of LGE suggests that access to all four Dixon phases, analogous to a human reader cross-referencing them, may help the model separate scar from adjacent EAT. These findings indicate that training and evaluating models on data reflecting real-world clinical variability may yield a more realistic estimate of generalizability than highly curated datasets.

### Segmentation Efficiency and Correction Workload

Under the intraobserver-derived threshold, only two of 30 test cases were flagged for manual review. Both occurred in contrast-enhanced studies of patients with underlying cardiovascular disease, which is consistent with segmentation complexity scaling with the presence of pronounced structural pathology. The number is too small to support formal conclusions, but the pattern fits a workflow in which a small subset of technically demanding cases continues to benefit from targeted manual quality control while the large majority of cases passes automated inference without intervention.

The time gain over manual segmentation is equally relevant for how EAT can be studied. Manual volumetric EAT quantification on isotropic 3D Dixon CMR takes around an hour per case, which has been a principal obstacle to integrating this approach into larger studies. Automated inference shifts this cost to one minute per case, enabling EAT assessment in large cohort sizes, multi-center datasets, and serial acquisitions that would not be feasible under manual segmentation.

### Comparison with Other EAT Quantification Approaches

EAT on CMR has often been quantified by manual contouring on two-dimensional short-axis cine stacks [4]. More recently, automated EAT quantification from CMR has been performed using different post-processing approaches, each shaped by the type of imaging available. The UK Biobank pipeline applies a neural network to a single four-chamber cine image to derive the pericardial adipose tissue area, which fits the 2D cine datasets available in this cohort [26]. Most isotropic work for EAT quantification has been performed using CT, where deep learning has shown good reproducibility and clear links to cardiovascular outcomes in large populations [27]. Isotropic 3D Dixon CMR offers a complementary option, with direct fat–water separation and no radiation exposure, rendering it suitable for serial and observational research.

For isotropic 3D Dixon imaging, the main practical constraint has been the time required for manual slice-by-slice segmentation. A pragmatic workaround has been to reslice the 3D dataset into two-dimensional short-axis stacks, reducing manual effort while keeping the segmentation format familiar [28]. Recent advances in deep learning–based segmentation offer an effective approach to overcome this post-processing limitation, enabling fast automated analysis of the full 3D dataset without compromising anatomical precision [16]. The automated model presented here operates directly on the full isotropic volume, maintaining spatial resolution while at the same time eliminating prolonged post-processing times. Feng et al. recently reported a shape-aware deep learning framework for EAT segmentation on 3D Dixon CMR, developed in patients with type 2 diabetes and matched controls and embedded in a broader morphogeometric analysis [29]. In contrast, the presented study uses the widely adopted 3D nnU-Net architecture and evaluates segmentation performance across healthy individuals and patients with cardiovascular disease, across low and high EAT burden subgroups, and in the presence of pathological imaging features and variable image quality.

### Possible Clinical and Research Implications

By making volumetric EAT quantification fast and reproducible, automated segmentation could facilitate the integration of EAT as an imaging biomarker into routine CMR workflows and large-scale studies. Increased EAT volume has been linked to adverse cardiac remodeling and cardiovascular events, yet its routine assessment has been limited by the post-processing burden of manual segmentation. The elimination of inter-observer offset is particularly relevant for longitudinal and multi-center assessment. As EAT can be reduced by cardiometabolic drug therapy, such as glucagon-like peptide-1 receptor agonists and sodium-glucose cotransporter-2 inhibitors, a fast and reproducible measurement could support monitoring of treatment-induced changes that would not be practical to track under manual segmentation [30]. While the present study focused on segmentation performance rather than clinical outcomes, it provides a technical foundation for subsequent studies that relate EAT volume measurements to cardiovascular outcomes and assess their value for risk stratification and treatment monitoring.

## Limitations

Despite covering different cardiac diseases, it was not possible to include all entities in this feasibility study. Cases with congenital heart disease were absent and coronary artery disease was underrepresented. Imaging was performed on two scanners at 1.5 T and 3 T, all from a single vendor.

## Conclusion

The automated segmentation model achieves agreement with manual reference segmentations comparable to intraobserver reproducibility for ventricular EAT quantification on isotropic 3D Dixon CMR. It performs consistently across healthy individuals and patients with underlying cardiovascular disease, across low and high EAT burden, and in the presence of pathological imaging features and variable image quality. Post-processing time is highly reduced from around an hour per case to one minute.

## Statements and Declarations

### Author Contribution

**HN:** Conceptualization, Data curation, Formal analysis, Funding acquisition, Investigation, Methodology, Project Administration, Software, Validation, Visualization, Writing – original draft, Writing – review & editing **RH:** Conceptualization, Data curation, Formal analysis, Funding acquisition, Methodology, Software, Supervision, Validation, Visualization, Writing – original draft, Writing – review & editing **CA:** Methodology, Writing – review & editing **JK:** Data curation, Investigation, Writing – review & editing **MF:** Data curation, Investigation, Writing – review & editing **CP:** Methodology, Writing – review & editing **RMB:** Methodology, Writing – review & editing **TH:** Methodology, Writing – review & editing **CH:** Software, Writing – review & editing **ED:** Writing – review & editing **EB:** Data curation, Investigation, Writing – review & editing **JG:** Writing – review & editing **CL:** Data curation, Writing – review & editing **JSM:** Conceptualization, Data curation, Funding acquisition, Investigation, Project administration, Resources, Supervision, Writing – original draft, Writing – review & editing

### Data Availability

The imaging data analyzed in this study are not publicly available due to patient privacy regulations and German legislation but may be made available by the corresponding author upon reasonable request.

### Code Availability

The trained model weights and inference pipeline are provided in this GitHub repository https://github.com/HalilNoyan/FatNet (available upon publication).

### Declaration of Generative AI Use

During the preparation of this manuscript, the authors used ChatGPT (OpenAI) for grammar checking and language editing. The authors reviewed and edited all AI-generated content and take full responsibility for the content of the published article.

## Data Availability

Data Availability
The imaging data analyzed in this study are not publicly available due to patient privacy regulations and German legislation but may be made available by the corresponding author upon reasonable request.
Code Availability
The trained model weights and inference pipeline are provided in the GitHub repository (available upon final publication).

https://github.com/HalilNoyan/FatNet

## Abbreviations

2CV: two-chamber view
2D: two-dimensional
3D: three-dimensional
4CV: four-chamber view
AI: artificial intelligence
ARVC: arrhythmogenic right ventricular cardiomyopathy
AV: atrioventricular
BMI: body mass index
BSA: body surface area
CAD: coronary artery disease
CI: confidence interval
CMR: cardiovascular magnetic resonance
CT: computed tomography
DSC: Dice similarity coefficient
EAT: epicardial adipose tissue
ECG: electrocardiogram
FP: fat-phase
GPU: graphics processing unit
HCM: hypertrophic cardiomyopathy
HD95: 95th percentile Hausdorff distance
IBD: inflammatory bowel disease
ICC: intraclass correlation coefficient
IP: in-phase
IQR: interquartile range
KDE: kernel density estimation
LGE: late gadolinium enhancement
LoA: limits of agreement
LVEF: left ventricular ejection fraction
MD: muscular dystrophy
MRI: magnetic resonance imaging
nnU-Net: no-new-U-Net (self-configuring deep-learning segmentation framework)
OP: opposed-phase
RVEF: right ventricular ejection fraction
RVOT: right ventricular outflow tract
SAX: short-axis
SCMR: Society for Cardiovascular Magnetic Resonance
SD: standard deviation
T: Tesla
TE1: first echo time
TE2: second echo time
VRAM: video random-access memory
WP: water-phase

## Supplementary

**Supplementary Table 1:** Diagnoses in the Dataset Splits. Values are presented as n (%). ARVC = arrhythmogenic right ventricular cardiomyopathy; CAD = Coronary Artery Disease; HCM = hypertrophic cardiomyopathy; IBD = inflammatory bowel disease; MD = muscular dystrophy; RVOT = right ventricular outflow tract.

| Diagnosis | Total Cases<br>(n = 195) | Train Set<br>(n = 165) | Test Set<br>(n = 30) |
| --- | --- | --- | --- |
| Healthy | 33 (16.9%) | 23 (13.9%) | 10 (33.3%) |
| Paroxysmal Atrial Fibrillation | 40 (20.5%) | 37 (22.4%) | 3 (10.0%) |
| CAD | 12 (6.2%) | 11 (6.7%) | 1 (3.3%) |
| HCM | 31 (15.9%) | 28 (17.0%) | 3 (10.0%) |
| IBD | 33 (16.9%) | 30 (18.2%) | 3 (10.0%) |
| MD | 38 (19.5%) | 35 (21.2%) | 3 (10.0%) |
| ARVC | 6 (3.1%) | 3 (1.8%) | 3 (10.0%) |
| Borderline ARVC | 7 (3.6%) | 4 (2.4%) | 3 (10.0%) |
| Biventricular ARVC | 2 (1.0%) | 2 (1.2%) | 0 (0.0%) |
| RVOT Tachycardia | 3 (1.5%) | 1 (0.6%) | 2 (6.7%) |
| Sarcoidosis | 2 (1.0%) | 2 (1.2%) | 0 (0.0%) |

## Detailed Training and Configuration of nnU-Net

### Network Architecture

A 3D nnU-Net (nnUNetv2, Version 2.6.2) was trained using the automatically configured 3d_fullres setup, resulting in a six-stage 3D PlainConvUNet. The network comprised 3D convolutional layers with a kernel size of 3 × 3 × 3 and two convolutional blocks per encoder and decoder stage. Feature map counts per stage were 32, 64, 128, 256, 320, and 320 channels. Each convolution was followed by instance normalization and LeakyReLU activation.

### Preprocessing

All images were resampled to an isotropic target spacing of 1.3 × 1.3 × 1.3 mm^3^, if necessary. Image data were resampled using third-order interpolation; segmentation masks using first-order interpolation. Images were cropped to non-zero foreground regions and normalized using channel-wise Z-score normalization.

### Training

Patch-based training used patches of 80 × 192 × 160 voxels with a batch size of 2. On-the-fly augmentation included spatial transformations (random rotations, scaling, elastic deformations, mirroring) and intensity-based augmentations. A five-fold cross-validation was conducted on 165 cases (132 training / 33 validation per fold), with case-level splits reused consistently across folds. Optimization used stochastic gradient descent with polynomial learning rate decay and a combined Dice and Cross-Entropy loss.

### Inference

Softmax probabilities from the five cross-validation models were averaged and converted to final segmentations via voxel-wise argmax classification.

### Computational Environment

Training was performed on a Linux workstation (Ubuntu 24.04.3) using PyTorch (Version 2.9.1) with CUDA 12.0 acceleration on an NVIDIA RTX 4000 Ada Generation GPU (20 GB of VRAM).

---

**Figure S 1.**
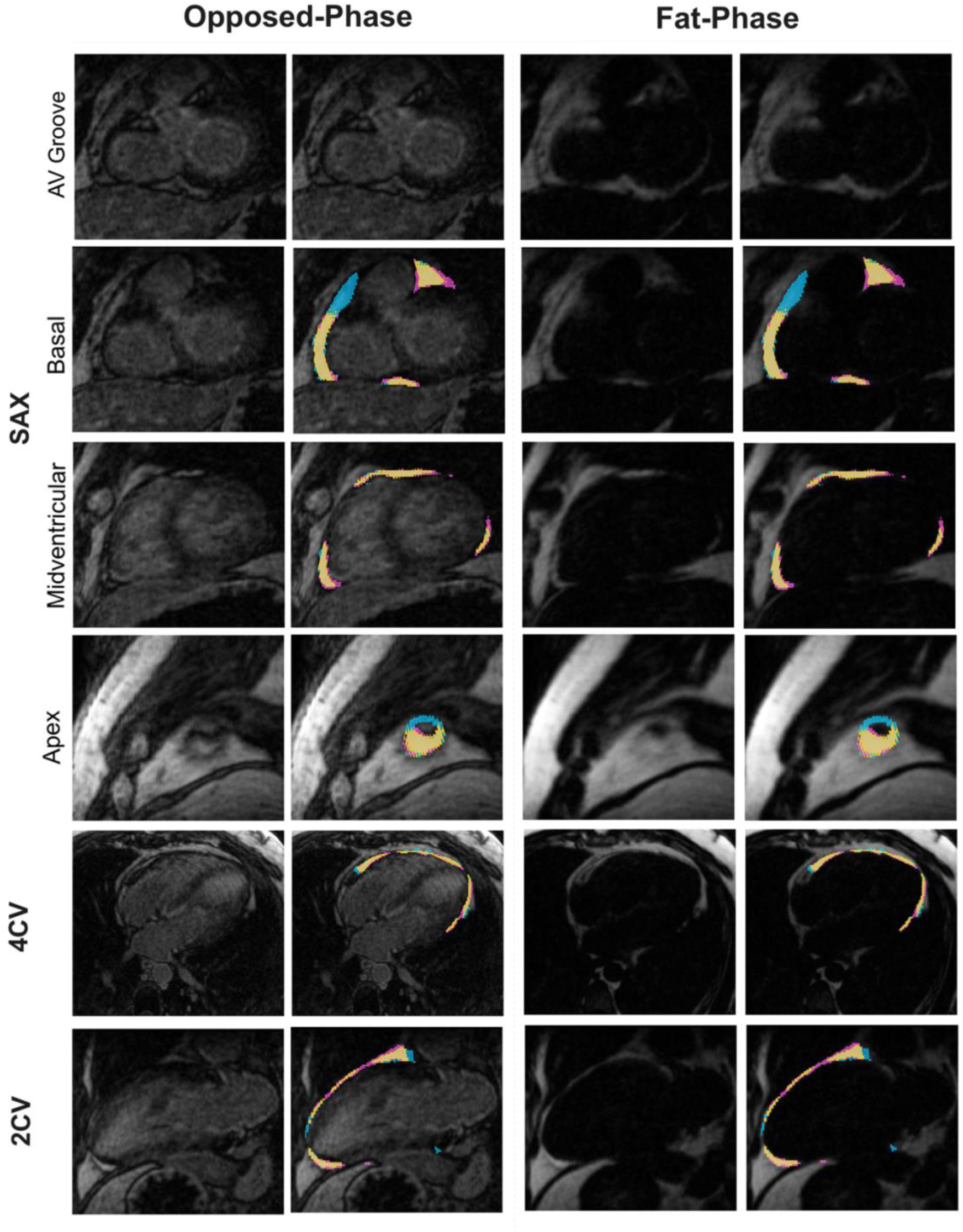
Qualitative comparison of automated EAT segmentation against manual ground truth for the flagged borderline arrhythmogenic right ventricular cardiomyopathy (ARVC) case across standard cardiac views. Each row shows a different view (SAX at atrioventricular (AV) groove, basal, midventricular, and apical levels; 4CV: four chamber view; 2CV: two chamber view) in opposed-phase and fat-phase images. Segmentation overlays (2nd and 4th column) are shown next to the corresponding source image (1st and 3rd column). Blue overlays indicate manual ground truth, magenta indicates model prediction, and orange indicates voxel-wise agreement between the two.

**Figure S 2.**
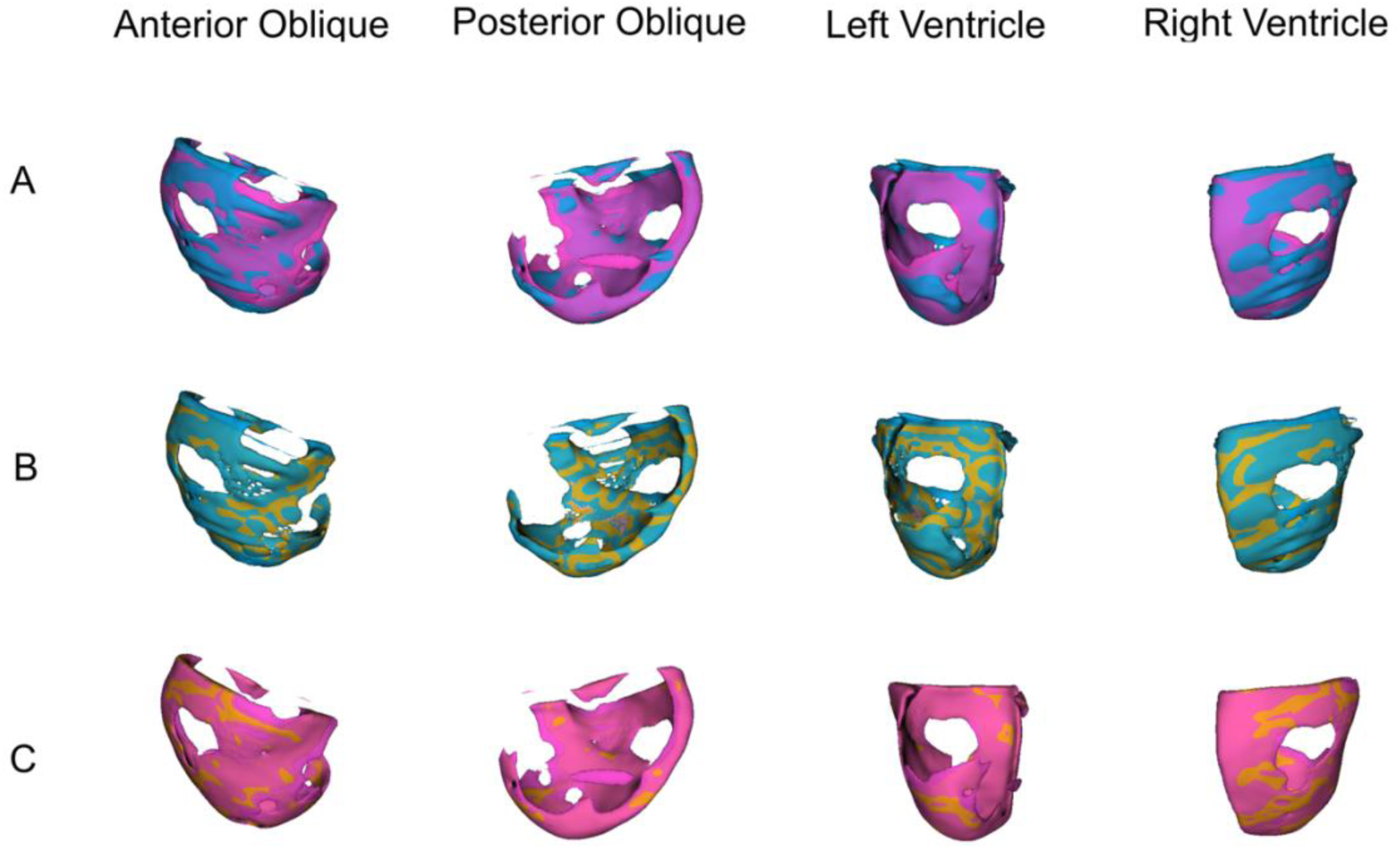
Three-dimensional renderings of epicardial adipose tissue (EAT) segmentation for the flagged borderline arrhythmogenic right ventricular cardiomyopathy (ARVC) case, shown from anterior-oblique and posterior-oblique perspectives, as well as from views facing the left-and right-ventricular sides of the heart. Each row shows a different pair of components to facilitate visual interpretation: (A) ground truth (blue) and model prediction (magenta); (B) ground truth (blue) and voxel-wise agreement (orange); (C) model prediction (magenta) and voxel-wise agreement (orange).

**Figure S 3.**
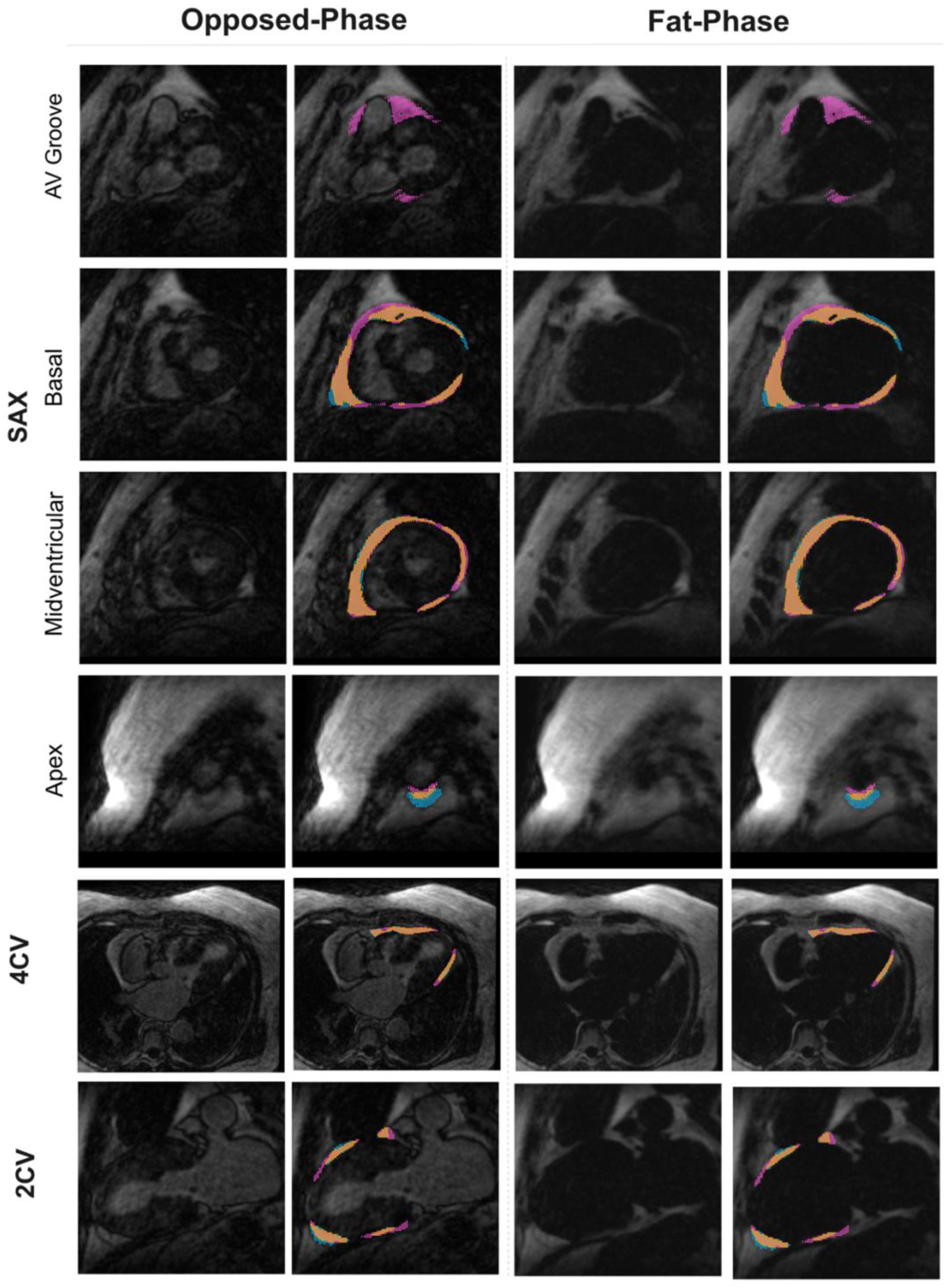
Qualitative comparison of automated EAT segmentation against manual ground truth for the flagged hypertrophic cardiomyopathy (HCM) case across standard cardiac views. Each row shows a different view (SAX at atrioventricular (AV) groove, basal, midventricular, and apical levels; 4CV: four chamber view; 2CV: two chamber view) in opposed-phase and fat-phase images. Segmentation overlays (2nd and 4th column) are shown next to the corresponding source image (1st and 3rd column). Blue overlays indicate manual ground truth, magenta indicates model prediction, and orange indicates voxel-wise agreement between the two.

**Figure S 4.**
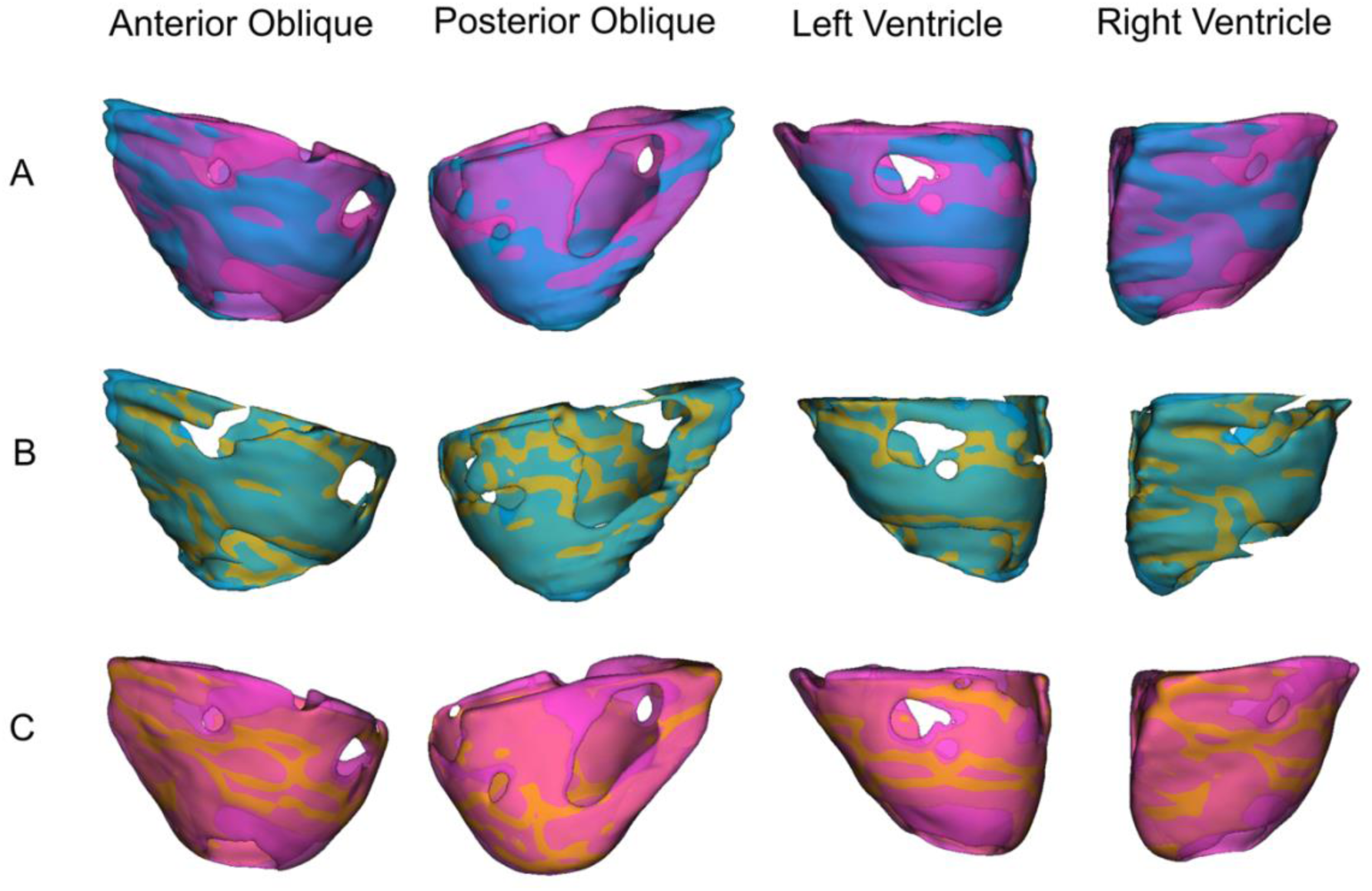
Three-dimensional renderings of epicardial adipose tissue (EAT) segmentation for the flagged hypertrophic cardiomyopathy (HCM) case, shown from anterior-oblique and posterior-oblique perspectives, as well as from views facing the left-and right-ventricular sides of the heart. Each row shows a different pair of components to facilitate visual interpretation: (A) ground truth (blue) and model prediction (magenta); (B) ground truth (blue) and voxel-wise agreement (orange); (C) model prediction (magenta) and voxel-wise agreement (orange).

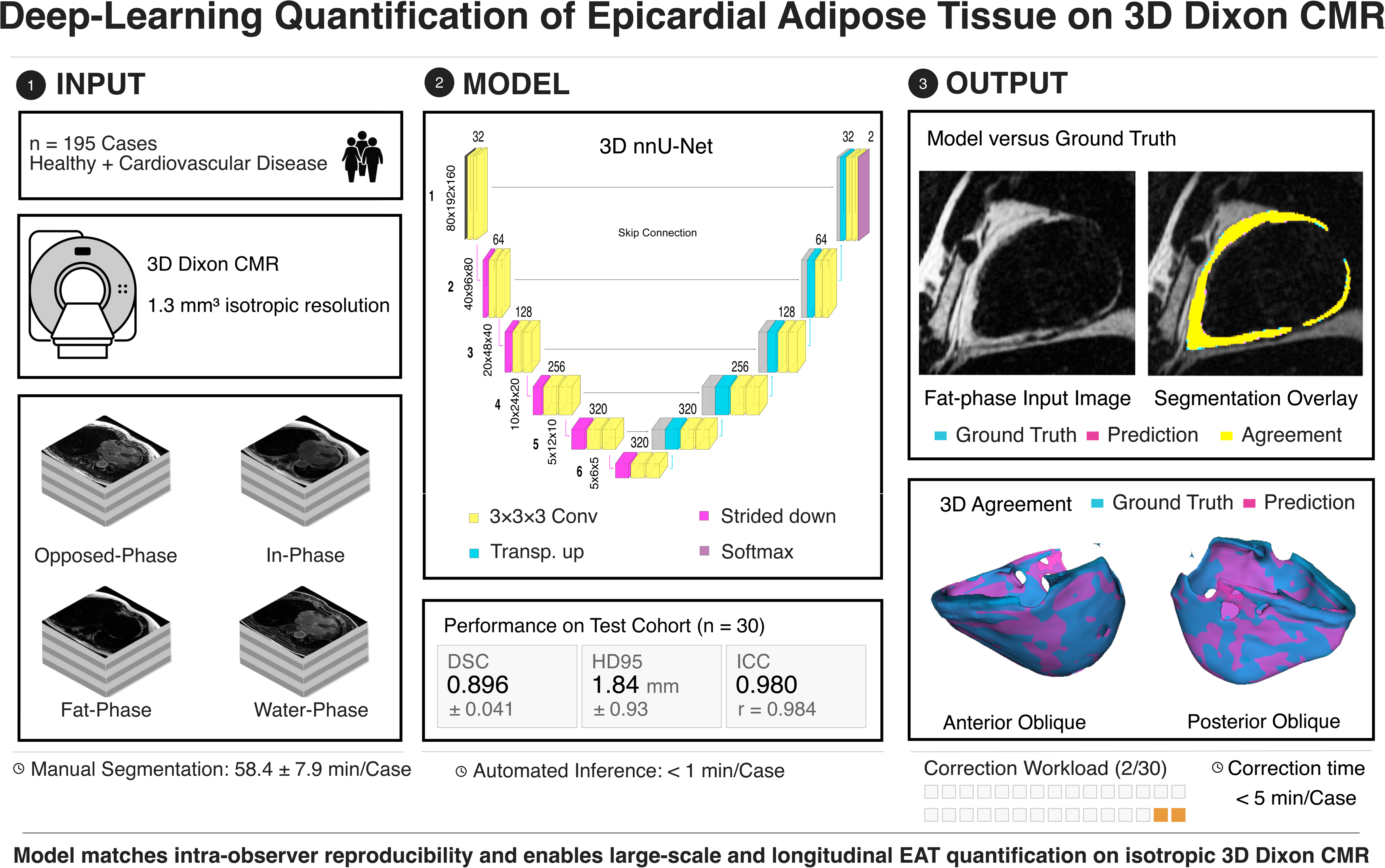

